# Performance of three molecular tests for SARS-CoV-2 on a university campus estimated jointly with Bayesian latent class modeling

**DOI:** 10.1101/2021.07.31.21261425

**Authors:** T. Alex Perkins, Melissa Stephens, Wendy Alvarez Barrios, Sean Cavany, Liz Rulli, Michael E. Pfrender

## Abstract

Accurate tests for severe acute respiratory syndrome coronavirus 2 (SARS-CoV-2) have been critical in efforts to control its spread. The accuracy of molecular tests for SARS-CoV-2 has been assessed numerous times, usually in reference to a gold standard diagnosis. One major disadvantage of that approach is the possibility of error due to inaccuracy of the gold standard, which is especially problematic for evaluating testing in a real-world surveillance context. We used an alternative approach known as Bayesian latent class modeling (BLCM), which circumvents the need to designate a gold standard by simultaneously estimating the accuracy of multiple tests. We applied this technique to a collection of 1,716 tests of three types applied to 853 individuals on a university campus during a one-week period in October 2020. We found that reverse transcriptase polymerase chain reaction (RT-PCR) testing of saliva samples performed at a campus facility had higher sensitivity (median: 0.923; 95% credible interval: 0.732-0.996) than RT-PCR testing of nasal samples performed at a commercial facility (median: 0.859; 95% CrI: 0.547-0.994). The reverse was true for specificity, although the specificity of saliva testing was still very high (median: 0.993; 95% CrI: 0.983-0.999). An antigen test was less sensitive and specific than both of the RT-PCR tests. These results suggest that RT-PCR testing of saliva samples at a campus facility can be an effective basis for surveillance screening to prevent SARS-CoV-2 transmission in a university setting.

## INTRODUCTION

Molecular testing has played a vital role in efforts to suppress transmission of SARS-CoV-2. This applies in both community settings (1, 2) and in more specialized settings, such as hospitals (3, 4), workplaces (5, 6), schools (7, 8), and travel (9, 10). Although many contextual factors affect the success of testing (11, 12), the foundation of any successful testing program is the availability of tests that are sufficiently sensitive and specific to achieve the program’s objectives.

Most evaluations of the sensitivity and specificity of molecular tests for SARS-CoV-2 have been performed in reference to a diagnostic that was considered a gold standard (13). Designating a diagnostic as a gold standard makes the calculation of sensitivity and specificity straightforward, as true positive (TP), true negative (TN), false positive (FP), and false negative (FN) test outcomes can all be defined clearly in reference to the gold standard. Under this assumption, sensitivity can be estimated as TP / (TP + FN) and specificity as TN / (TN + FP).

A key limitation of this approach is that the estimates it yields are only as reliable as the gold standard on which they are based. The most common gold standard is reverse transcriptase polymerase chain reaction (RT-PCR) testing (14). This standard is far from golden, however. Especially with respect to sensitivity, the performance of these tests for SARS-CoV-2 has been found to vary as a function of the method of sample extraction (15, 16), day of infection (17, 18), and disease severity of the subject (19). Furthermore, designation of one method as a gold standard makes it impossible to evaluate whether another test might actually have better sensitivity or specificity than the presumed gold standard (20).

One way to circumvent the limitations associated with relying on a gold standard is to use an alternative method for analysis, such as Bayesian latent class modeling (BLCM) (21). This method involves joint estimation of the sensitivity and specificity of each type of test used, by virtue of considering the possibility that any given test result could have been erroneous for some, all, or none of the tests used. This approach has been applied in some cases for molecular tests for SARS-CoV-2, resulting in differences relative to estimates that relied on a gold standard (22–24). For example, in a meta-analysis comparing RT-PCR testing of nasopharyngeal and saliva samples, allowing for imperfections in both types of tests resulted in higher estimates of specificity and narrower uncertainty about sensitivity (23).

In this study, we applied BLCM to a data set from a SARS-CoV-2 testing program in a university setting during October 2020. A unique feature of this data set is that it includes both RT-PCR and antigen tests, which have not been compared in previous BLCM analyses for SARS-CoV-2 that we are aware of (22–24). Another unique feature of this data set is that the majority of subjects were tested for surveillance screening and were not suspected of being infected at the time of testing. This presents an opportunity to quantify test performance in a context that is highly relevant for public health (11). Moreover, the fact that the majority of subjects were in the 18-25 age range presents an opportunity to quantify test performance in a population for which tests may be less sensitive (19) yet are of high value for surveillance (7, 8).

## METHODS

### Sample collection

All samples for this study were collected during a five-day period from Monday, October 12 through Friday, October 16, 2020. In total, 1,716 tests were performed on samples collected from 853 individuals, with multiple tests for a single individual applied to specimens collected on the same day. Most individuals (811) participated in response to a request for surveillance testing, while others (42) participated either as a result of reporting symptoms associated with COVID-19 (29), because of suspected exposure through contact (10), or because they had previously tested positive and were undergoing a second test four days later (3). Participants consisted of 846 students, and 7 faculty and staff. The majority (87.6%) of students were between the ages of 18 and 22 inclusive, with a range of 18-39, a median age of 20, and a mean age of 21.2. The median age of staff was 40, and two members of staff were over the age of 65, with a range of 29-72 (Fig. S1). These individuals received a total of 833 commercial

RT-PCR tests on nasal swab specimens, 846 in-house RT-PCR tests on saliva specimens, and 37 antigen tests on nasal swab specimens. We refer to these tests hereafter as commercial, saliva, and antigen tests, respectively. A majority of individuals (799) received commercial and saliva tests but not an antigen test, a subset (27) received all three tests, and the remainder (27) received either one or two tests in other combinations.

### Laboratory testing

#### SARS-CoV-2 Detection in Saliva Samples

Following the University of Notre Dame IBC approved protocol (20-08-6161), fresh saliva samples were obtained from study participants and tested for the presence of SARS-CoV-2 within 17 hours after collection. Steps for the detection of SARS-CoV-2 through RT-qPCR in saliva samples were adapted from Ranoa et al. (25). A minimum of 200 μl of saliva were collected from each participant in a barcoded nuclease free 50mL conical tube. Following collection, samples were heat inactivated by incubating in a 95 °C circulating water bath for 30 min. After cooling to room temperature, the inactive specimen was diluted at a 1:1 ratio (vol/vol) with 2xTris-Borate-EDTA buffer (0.089M Tris, 0.089M Borate, 0.002M EDTA in final 1x buffer solution), followed by vigorous vortexing to ensure thorough mixing. The diluted saliva was then subjected to RT-qPCR (1 reaction per sample) using the TaqPath COVID-19 Combo kit (Thermo Fisher Scientific), which includes three primer/probe sets specific to SARS-CoV-2 genes (ORF1ab, S-gene, N-gene), and one MS2 bacteriophage control target. Briefly, 5μl diluted saliva was added to a freshly prepared reaction mix containing 2.5μl TaqPath1-Step Multiplex Master Mix (No ROX) (4X), 0.5μl COVID-19 Real Time PCR Assay Multiplex, 0.5μl MS2 phage control, and 1.5μl nuclease-free water. Reactions were set up in a 96-well plate format (0.1mL MicroAmp Fast Optical 96-Well reaction plate (Applied Biosystems)), with each plate containing a positive control diluted to 4 copies/μl and a no template control (nuclease-free water), as well as the MS2 control internal to each sample. All RT-qPCR reactions were carried out using QuantStudio RT-qPCR instruments (Applied Biosystems). Reaction parameters were set as follows: *Hold Stage* 25 °C 2:00min, 53 °C 10:00min, 95 °C 2:00min; *PCR Stage* (40X) 95 °C 3sec, 60 °C 30sec; 1.6 °C/sec ramp for all stages; run mode “fast.” Targets, reporter dyes, and quencher information for RT-qPCR instrument was set up according to TaqPath COVID-19 Combo kit manufacturer’s instructions.

Presence/Absence analysis of the viral targets was performed using Applied Biosystems Design and Analysis v2.4 software with a baseline set at 5 and Cq cutoff for all targets set at 37. Results of the RT-qPCR test were interpreted as positive, negative, or invalid. A positive test had at least 2 of the 3 gene targets present within the threshold settings. All positive and invalid tests were subjected to repeat testing for confirmation.

#### SARS-CoV-2 Commercial and Rapid Antigen Detection

Self-administered nasal swab samples were outsourced to LabCorp Inc. for viral detection with a RT-PCR protocol (EUA200011). Rapid antigen assays were performed on self-administered nasal swab samples with the Sofia2 Fluorescent Immunoassay Analyzer and the SARS Antigen Fluorescent Immunoassay (FIA) for qualitative detection of the nucleocapsid protein from SARS-CoV-2 (Quidel).

### Statistical analysis

#### Model

For our analysis, we estimated eight parameters (Table 1), which together determine the probability of each type of testing outcome. The likelihood of a given set of values of these parameters is equal to the probability of the observed testing outcomes given those parameter values. The data were defined according to the number of individuals with a given combination of testing outcomes as *n*_*i,j,k*_, where *i, j*, and *k* refer to positive, negative, or missing results for each of commercial, saliva, and antigen tests, respectively (Table 2).

**Table 1.**
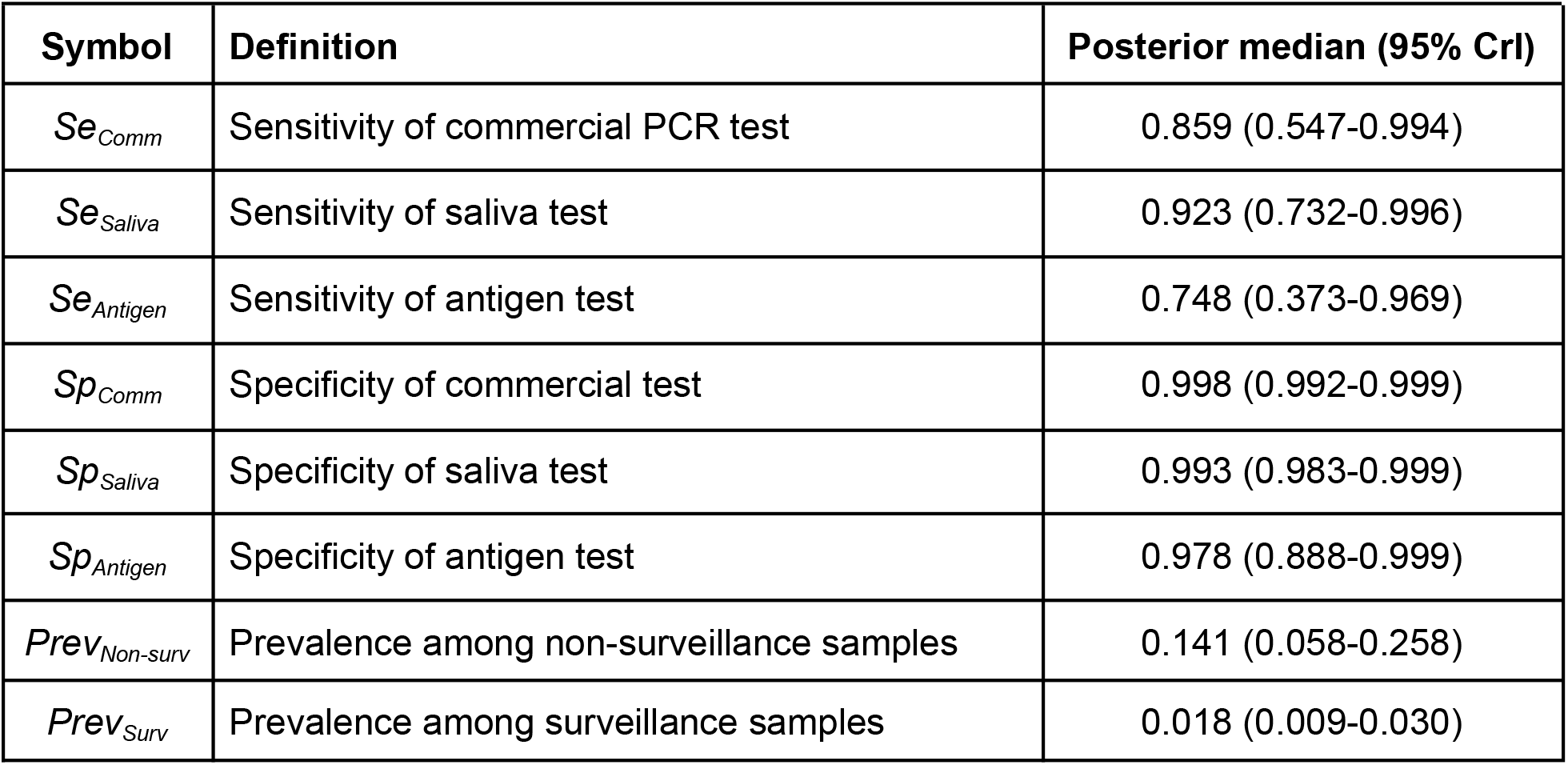
Parameter definitions and posterior estimates.

**Table 2.** Testing data. Each of the 853 study participants fell into one of the categories represented by each row. These categories differed with respect to the reason for testing and the outcome of each test. NA indicates that a given test was not performed for those individuals. This table constitutes the full information used in our analysis.

| Reason for testing | Commercial | Saliva | Antigen | Number of participants |
| --- | --- | --- | --- | --- |
| Surveillance | - | - | - | 3 |
|  | - | - | NA | 778 |
|  | - | NA | NA | 7 |
|  | NA | - | NA | 3 |
|  | - | + | NA | 4 |
|  | + | - | NA | 1 |
|  | + | + | NA | 9 |
|  | NA | + | NA | 7 |
| Other | - | - | - | 22 |
|  | - | - | NA | 7 |
|  | NA | - | - | 5 |
|  | NA | + | + | 4 |

By definition, test sensitivity and specificity are specified in reference to the true infection status of an individual. Because we did not know the true infection status of any individual with certainty, we defined the probability of a given set of testing outcomes (i.e., *i, j, k*) conditional on the true status, which we refer to as *s* (this could be either + or -). This probability, Pr(*i,j,k*|*s*), is defined as the product of the probabilities of each testing outcome given status *s*.

To account for the fact that the true status of any given infection is unknown, we used the law of total probability to calculate the overall probability of the observed testing outcomes, Pr(*i,j,k*), as the weighted average of the conditional probabilities of the observed testing outcomes,

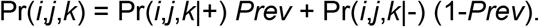

Denoting the set of all parameters as ***θ***, we defined the likelihood of the parameters given the data, ***n***, as

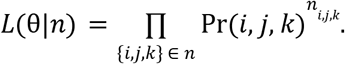

In these calculations, different values of *Prev* are used depending on whether the individuals were tested as part of surveillance efforts or for other reasons.

#### Estimation procedure

Taking a Bayesian approach to parameter estimation, the posterior probability of the parameters in ***θ*** was defined as

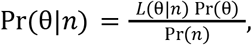

where L(***θ***|***n***) is the likelihood defined above, Pr(***θ***) is the prior probability of the parameters, and Pr(***n***) is the probability of the data. We assumed non-informative priors for sensitivity and specificity parameters, and we assumed informative priors for the two types of prevalence that were in loose alignment with estimated prevalence at the time and location of sample collection. We avoided calculation of Pr(***n***) by using Markov chain Monte Carlo (MCMC) sampling. Details about the prior assumptions and MCMC algorithm are provided in the Supplemental Text.

#### Validation

To validate our model, we applied it to 100 simulated data sets and compared inferred parameter values to the true parameter values used to simulate the data. To ensure that the model’s inferences were valid for data resembling those used in this study, we simulated the same number of individuals tested with the same combination of tests as in our empirical data. Simulated parameter values were drawn uniformly and independently from the 95% credible interval of each parameter. We examined coverage probabilities and correlations between median and true parameter values across the 100 simulated data sets.

#### Predictive value

Using the posterior parameter estimates, we calculated predictive values of the three tests under two different contexts. These values represent the probability that the test’s indication, whether positive or negative, reflects the true status of the individual being tested. The positive predictive value is defined as

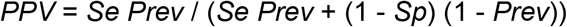

and the negative predictive value is defined as

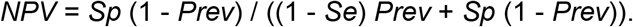

First, we calculated *PPV* and *NPV* during the one-week period of our study, accounting for uncertainty in *Prev* in doing so. Second, we calculated *PPV* and *NPV* on a daily basis over the course of the entire fall 2020 semester, accounting for daily changes in prevalence over time. Our estimates for time-varying prevalence were based on an extrapolation of the daily incidence of symptomatic cases (26) that accounted for the proportion symptomatic (27), the incubation period (28), and the probability that a test administered on a given day of infection would be positive (11). This method is described in further detail in the Supplemental Text.

## RESULTS

Pooling across all tests performed, test positivity was 2.5% (43/1,716). Positivity was lower among individuals tested for surveillance purposes (1.9%) than among individuals tested for other reasons (12.3%). Positivity was also lower among commercial tests (1.4%) than saliva (3.1%) and antigen (13.5%) tests. Lower positivity among commercial tests held when controlling for the method by which individuals came to participate in the study (Table S1). Despite differences in positivity, the very low positivity overall meant that concordance was high: 99.3% between commercial and saliva tests, 96.3% between commercial and antigen tests, and 97.3% between saliva and antigen tests.

Our Bayesian analysis leveraged joint information about all observed combinations of testing outcomes across the three types of tests (Table 2) to estimate a total of eight parameters (Table 1). Application of this method to 100 simulated data sets showed good coverage of true parameter values (Supplemental Text, Fig. S2). Applying the method to empirical data demonstrated good convergence (Fig. S3) and resulted in posterior samples with moderately low correlation (Fig. S4), suggesting that the data were reasonably informative about the parameters we sought to estimate.

Prevalence inferred by our Bayesian analysis was similar to test positivity for surveillance testing (median: 1.8%; 95% credible interval: 0.9-3.0%) (Fig. 1A) and slightly higher for non-surveillance testing (median: 14.1%; 95% CrI: 5.8-25.8%) (Fig. 1B). For saliva tests, we estimated a sensitivity of 0.923 (95% CrI: 0.732-0.996) (Fig. 1C, green) and a specificity of 0.993 (95% CrI: 0.983-0.999) (Fig. 1D, green). Had we considered commercial tests to be a gold standard, we would have instead estimated the sensitivity and specificity of saliva tests to be 0.833 and 0.995, respectively. Similarly, for antigen tests, our estimate of sensitivity (0.748; 95% CrI: 0.373-0.969) (Fig. 1C, blue) was greater than an estimate made in reference to commercial tests (0.5), and our estimate of specificity (0.978; 95% CrI: 0.888-0.999) (Fig. 1D, blue) was lower than an estimate made in reference to commercial tests (1.0). These discrepancies were a result of the fact that we did not consider commercial tests to be a gold standard and estimated their sensitivity and specificity alongside that of the other two test types. Doing so resulted in estimates of sensitivity of 0.859 (95% CrI: 0.547-0.994) (Fig. 1C, red) and specificity of 0.998 (95% CrI: 0.992-0.999) (Fig. 1D, red) for commercial tests.

**Figure 1.**
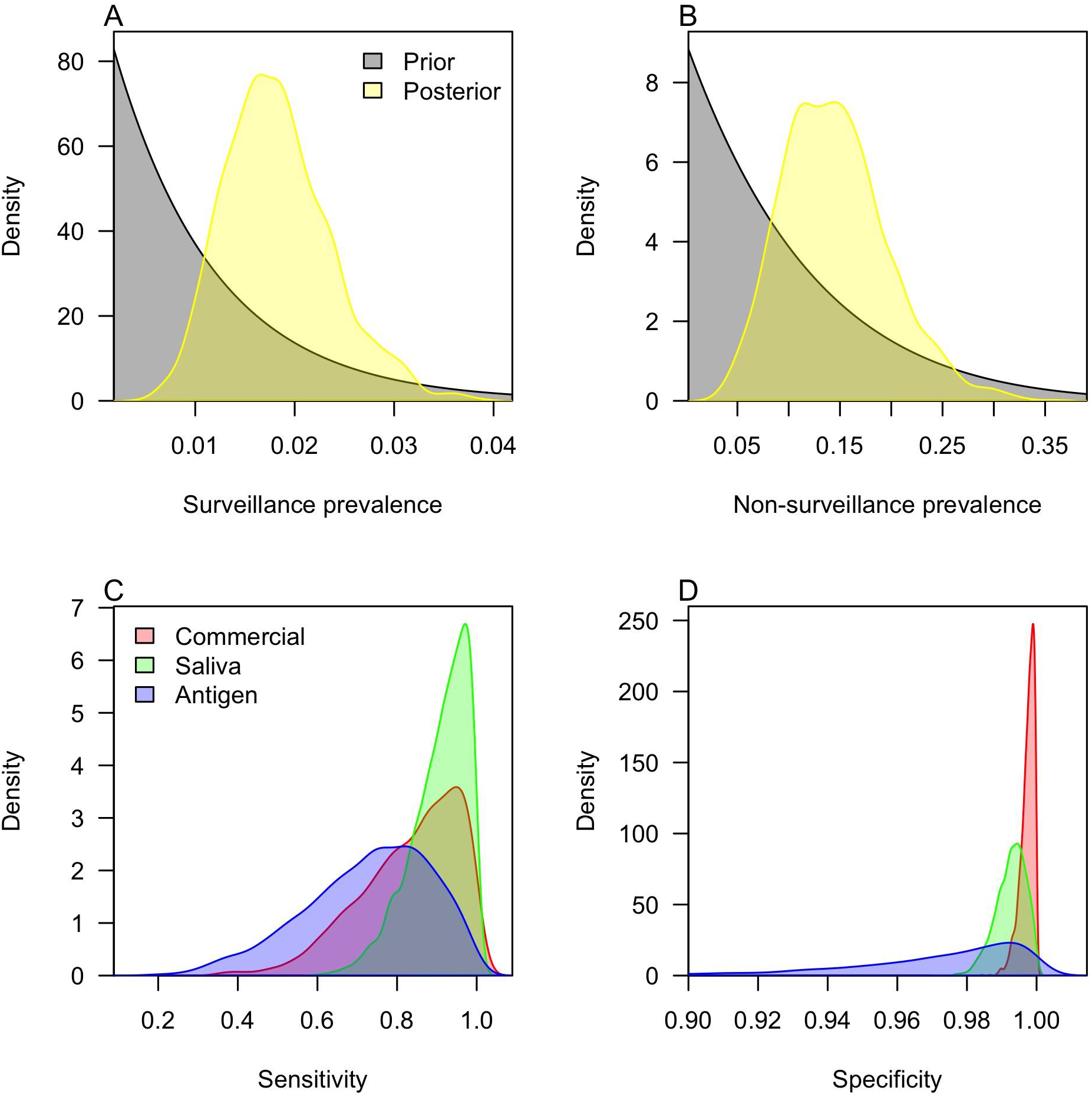
Posterior parameter estimates. (A) Prevalence among individuals participating in surveillance testing; (B) prevalence among individuals participating in testing for reasons other than surveillance; (C) test sensitivity; and (D) test specificity. Colors in A and B distinguish prior from posterior distributions, and colors in C and D distinguish different types of tests. Values outside the 0-1 range occur only as a result of smoothing.

While comparison of median sensitivities and specificities implies that some tests were more sensitive or specific than others, the wide uncertainty of our estimates must be considered when making such comparisons. We obtained more nuanced insight into the relative sensitivities and specificities of the three tests by calculating the proportion of samples in which the sensitivity of one test exceeded that of another, and likewise for specificity. On that basis, we found a probability of 0.69 that the saliva test was more sensitive than the commercial test (Table S2). The probabilities that the saliva and commercial tests were more sensitive than the antigen test were 0.88 and 0.71, respectively. The probabilities that the commercial test was more specific than the saliva and antigen tests were 0.86 and 0.92, respectively (Table S3). The saliva test was more specific than the antigen test with probability 0.81.

Joint inference of test properties and the prevalence of infection allowed us to estimate the frequency of different outcomes from surveillance testing (Fig. 2). Due to its high sensitivity, the saliva test was predicted to yield the most true positives (16.1 per 1,000 tests; 95% CrI: 8.2-27.6) (Fig. 2A) and the fewest false negatives (1.3 per 1,000 tests; 95% CrI: 0.07-5.4) (Fig. 3C). At the other extreme, 1,000 antigen tests were predicted to yield 12.7 true positives (95% CrI: 5.4-23.9) and 4.4 false negatives (95% CrI: 0.5-13.1). Antigen tests also had the lowest specificity, resulting in the largest number of false positives (21.3 per 1,000 tests; 95% CrI: 0.8-110.3) (Fig. 2B). Commercial tests were estimated to perform best in this regard, yielding only 2.1 false positives per 1,000 tests (95% CrI: 0.1-7.5).

**Figure 2.**
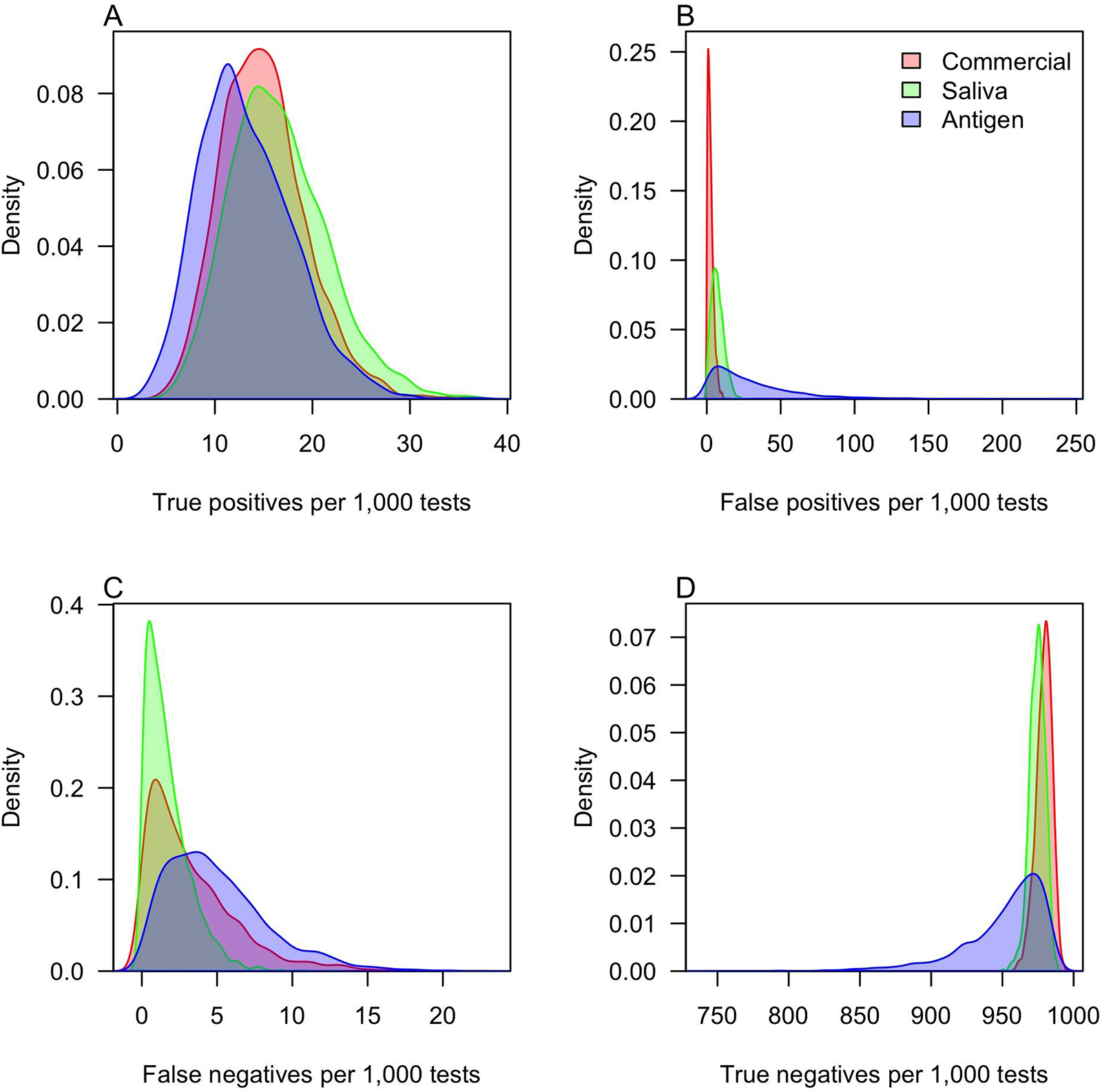
Estimates of the frequency of different testing outcomes. Out of 1,000 tests, panels show the number of (A) true positives, (B) false positives, (C) false negatives, and (D) true negatives. Colors distinguish different types of tests. Values outside the 0-1,000 range occur only as a result of smoothing.

**Figure 3.**
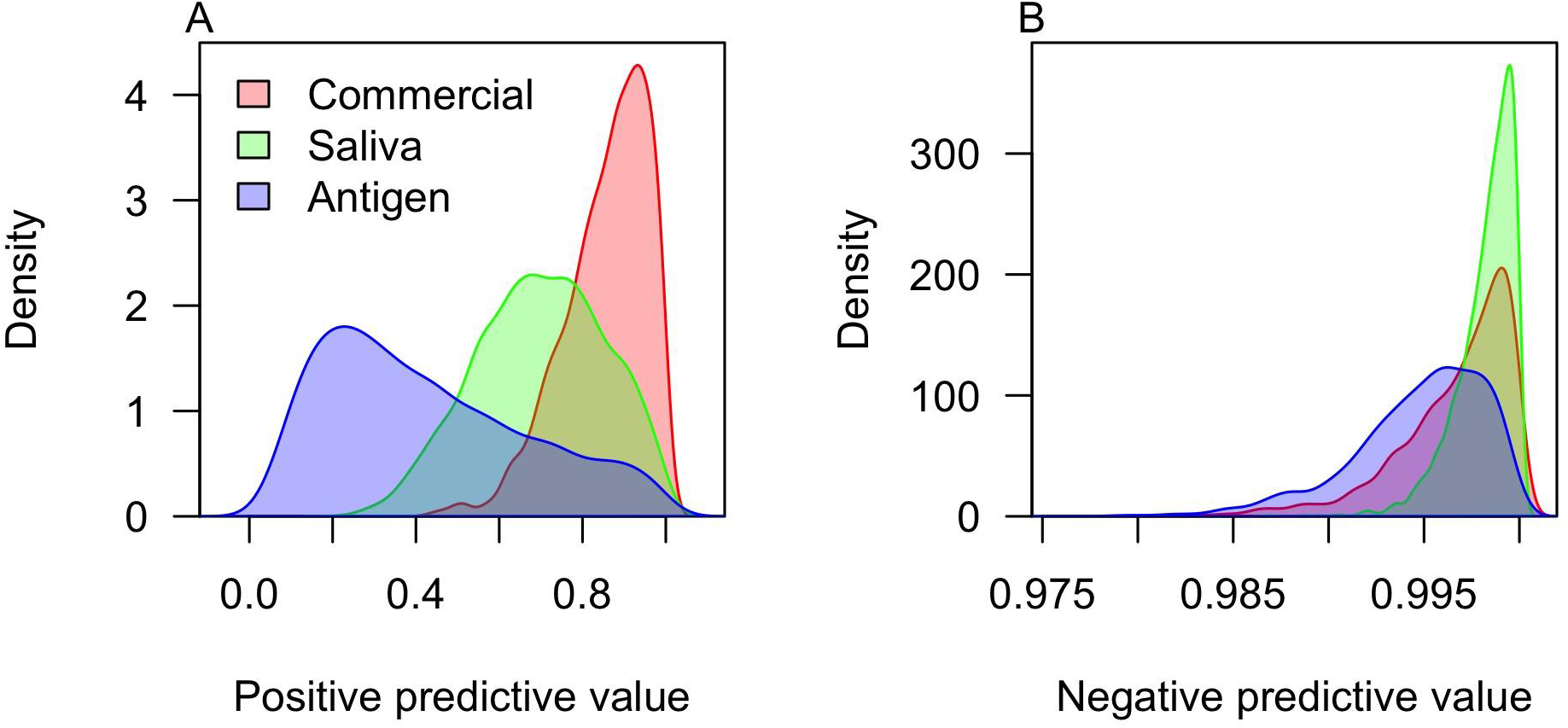
Estimates of the predictive values of each test during the study period. Panels show estimates of (A) the positive predictive value and (B) the negative predictive value. Colors distinguish different types of tests. Values outside the 0-1 range occur only as a result of smoothing.

Because of their high specificity and the low prevalence of infection, commercial tests had the highest positive predictive value during the study period (0.87; 95% CrI: 0.62-0.99) (Fig. 3A). Given such low prevalence, all tests had high negative predictive values and were predicted to result in mostly true negatives (Fig. 3B) for the vast majority of tests. Under a scenario of surveillance screening at random from the campus population during the fall 2020 semester, we estimated that saliva tests during the semester would be expected to have median positive predictive values as low as 0.001 (95% CrI: 0.0004-0.012) on August 1 and as high as 0.82 (95% CrI: 0.64-0.98) on August 22 (Fig. 4B). Negative predictive values of the saliva test never would have been less than a median of 0.997 (95% CrI: 0.990-0.999) under this scenario (Fig. 4E). Commercial tests would have had higher positive predictive values under this scenario (Fig. 4A), and both commercial and antigen tests would have had lower negative predictive values (Fig. 4D & F).

**Figure 4.**
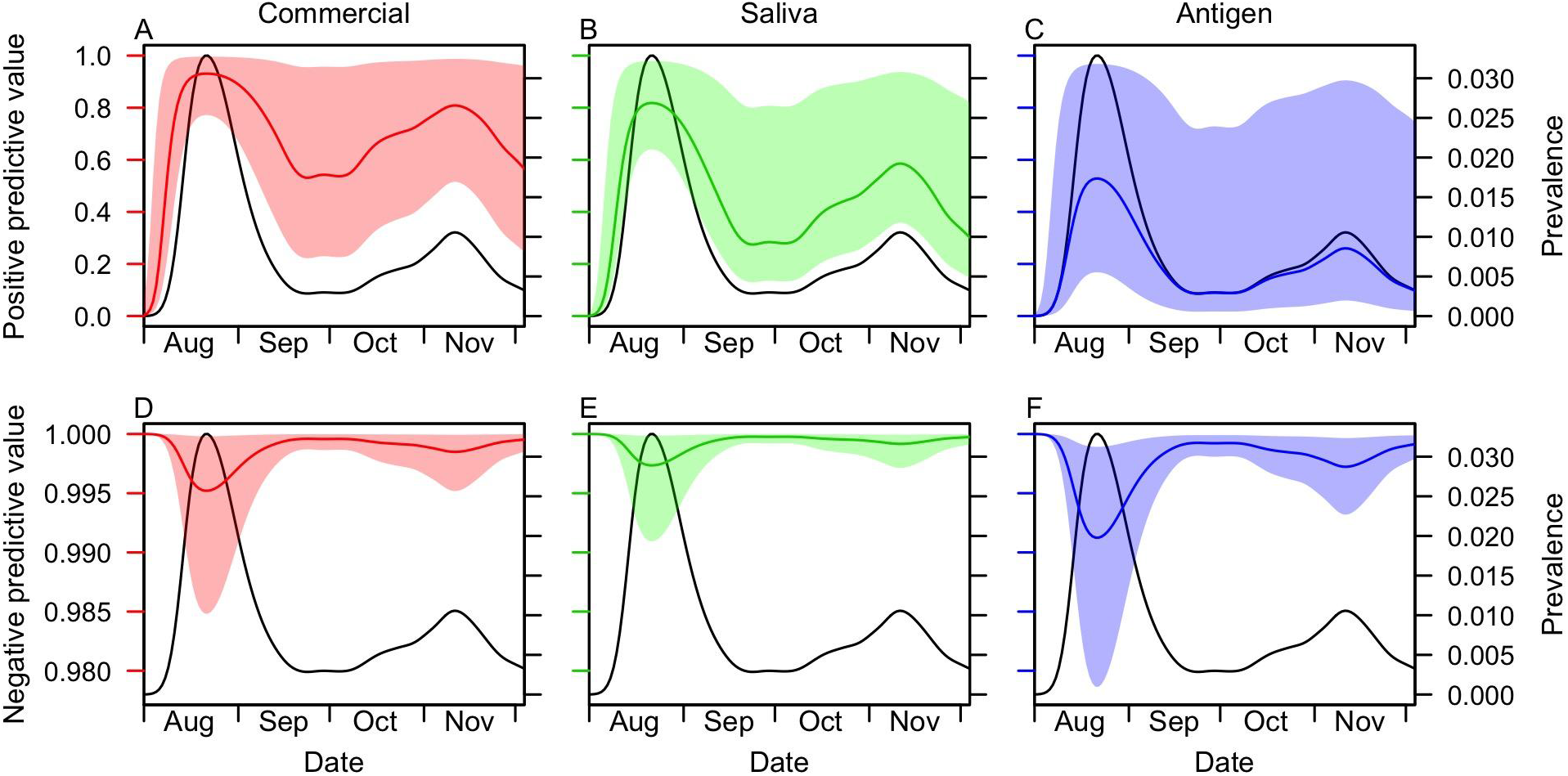
Positive predictive value (top) and negative predictive value (bottom) over the course of the entire semester. These values represent the probability that a positive or negative test result under random surveillance screening would have accurately relayed the true positive or negative status of the individual being tested. Change over time was a result of time-varying prevalence of detectable infection (black lines, right axis). Uncertainty reflects uncertainty about sensitivity and specificity of each type of test: commercial (red), saliva (green), antigen (blue).

## DISCUSSION

With respect to sensitivity, there were important differences between our modeled estimates and those based on raw test data. The clearest difference was for the commercial test, for which we obtained a median estimate of 86% for sensitivity. Had we considered that test to be a gold standard, we would have obtained a point estimate for the sensitivity of the saliva test 16% lower than our median estimate, and 33% lower for the antigen test. On the contrary, we found support for the saliva test likely being more sensitive than the commercial test, which would not have been possible to infer had we assumed a gold standard (20). Our finding that the saliva test was more sensitive than the commercial test, which was based on a nasal swab sample, is consistent with findings from other studies (29–32). We estimated that the antigen test had markedly lower sensitivity than either of the RT-PCR tests, which is also consistent with other studies (33). Given that only 37 individuals received an antigen test, it is important to bear in mind that uncertainty about its properties is high.

With respect to specificity, the medians of our modeled estimates were slightly lower than estimates based on the commercial test as a gold standard. Even so, our specificity estimate for the saliva test (median: 0.993; 95% CrI: 0.983-0.999) was strikingly similar to an independent estimate (median: 0.992; 95% CrI: 0.982-0.998) that also used a BLCM, but in the context of a meta-analysis (23). Our analysis generated high confidence in the commercial test being the most specific and the antigen test being the least specific, with probability 0.8. Our median estimate of 0.978 for antigen test specificity was within the range of published estimates for seven different antigen tests (34), which had median values ranging from 0.985 to 1.0 for five tests and from 0.889 to 0.948 for two outliers. Uncertainty about test specificity was relatively low in our estimates, given that the vast majority of the individuals we tested were likely true negatives.

These test sensitivities and specificities have implications for several metrics of public health importance. Given its high sensitivity, the saliva test was expected to detect the most true positives and produce the fewest false negatives, as indicated by a high negative predictive value. For the purpose of identifying infections in surveillance screening so that they can be isolated and their transmission curtailed, this test was most ideal, especially at times of high prevalence. Given its high specificity, the commercial test was expected to result in the fewest false positives and the most true negatives, as indicated by a high positive predictive value.

These properties are ideal from the perspective of minimizing unnecessary demand on resources for case isolation and contact tracing, more so at times of low prevalence when demand is already low. The antigen test performed least well in all regards. While this may make it seem like a less desirable option, it should be noted that sensitivity over the course of infection as a whole need not be paramount. In the event that a test has high sensitivity around the time of peak infectiousness, its value for curtailing transmission could still be very high (35).

Given the implications of our estimates of sensitivity and specificity, it is important to understand their empirical basis. It is notable that, out of 853 individuals tested, only six had discrepant results. In four of those cases, the saliva test was positive and the commercial test was negative. In two, the commercial test was positive and one or both of the other tests were negative. A strength of our modeling approach is that it integrated across all of the available information to inform its estimates, rather than those six discrepancies alone. The model also took into account the higher positivity of the saliva and antigen tests, as compared to the commercial test. Likewise, it was capable of balancing that with indications that the antigen test had lower specificity, which could explain its higher observed test positivity in part. Additionally, the model was able to account for higher positivity among non-surveillance tests due to higher prevalence in that group, which is important given that the three types of tests were not applied evenly across the two groups. These competing influences on our estimates underscore the value of the BLCM approach we used, which was able to balance them appropriately and express that balance in the form of quantitative descriptions of uncertainty.

Although our analysis was able to provide insight into the properties of these three tests, there were some uncertainties that we were unable to resolve. Correlations among three parameters—*Se*_*Comm*_, *Sp*_*Saliva*_, and *Prev*_*Surv*_—were indicative of uncertainty about whether the four individuals with positive saliva tests and negative commercial tests resulted from false-positive saliva tests or false-negative commercial tests. More data would be helpful for resolving this uncertainty, although doing so would require relatively rare discrepant results. Another limitation of our study is that the only information we used to resolve uncertainty about true infection status was whether individuals were tested for surveillance purposes. Given that prevalence differed by an order of magnitude between these groups, this was quite beneficial. However, additional information—such as recent contacts or status as student, faculty or staff—could have potentially helped narrow this type of uncertainty further. Doing so would have required estimating more parameters and could have made the analysis more susceptible to bias if an increasingly complex model were not specified properly.

In addition to limitations of our analysis, there were also limitations of our data set. First, a more balanced testing effort across different test types and groups of subjects could have helped reduce uncertainty about certain parameters, especially those relating to antigen tests. Second, for individuals who truly were positive, we have no information about how many days elapsed between their initial exposure and when they were tested. Given how variable test sensitivity is over the course of an infection (11, 17), this factor alone could be a major driver of the sensitivities we estimated. For example, individuals tested for surveillance screening had presumably not displayed any symptoms up to the time of testing, making it possible that many of our positive surveillance tests came from presymptomatic individuals with high viral loads (36). Even so, the balance of saliva and commercial tests across individuals tested for surveillance versus non-surveillance purposes was similar, meaning that any differences in the timing of testing between these two groups should not have affected our inferences about the relative sensitivities of these two types of tests.

In conclusion, our analysis leveraged all data collected from this study to estimate the sensitivities and specificities of three types of tests, without the need to consider any of those tests as a gold standard. These estimates are pertinent to a setting for which surveillance testing has been (37, 38), and remains (39), a major emphasis of COVID-19 prevention.

Although there is appreciable uncertainty associated with our estimates, this uncertainty was quantified carefully and could be reduced in the future by updating our estimates with additional data. Bayesian analyses lend themselves to this naturally, given that posterior estimates from one study can serve as prior estimates for another. All code and data from this study is available at https://github.com/TAlexPerkins/SARSCoV2_BLCM to facilitate applications or extensions of this work by others.

## Data Availability

All code and data from this study is available at https://github.com/TAlexPerkins/SARSCoV2_BLCM.

## ACKNOWLEDGEMENTS

We thank Paul Hergenrother for generously sharing his expertise and experience designing and implementing a saliva-based COVID-19 surveillance program at Illinois University, Champaign-Urbana. Special thanks to Joanna McNulty and Carol Mullaney for coordinating the sample collection efforts and the technicians in the Notre Dame COVID Surveillance Lab who assisted with sample processing and data collection. This study was made possible by the enthusiastic participation of Notre Dame undergraduate students, staff, and faculty. Funding was provided by the University of Notre Dame and Notre Dame Research. This study was conducted under University of Notre Dame IRB #21-06-6687.

## SUPPLEMENTAL TEXT

### Prior distributions

Regarding the prior probabilities of the parameters, Pr(***θ***), we assumed a uniform prior for all sensitivity and specificity parameters. We explored uniform priors for the prevalence parameters but found that such a choice could result in very high estimates of prevalence and very low estimates of sensitivity and specificity, which seemed implausible based on other studies (23, 40). To reflect our assumption that the tests are likely more accurate than not and that infection prevalence is likely to be relatively low, we adopted beta distributed priors with shape parameters 1 and 99 for individuals recruited for surveillance testing. These parameters correspond to a distribution with a mean infection prevalence of 0.01 skewed towards lower values, which is generally consistent with model-based estimates of infection prevalence for St. Joseph County, Indiana (40). For individuals tested for non-surveillance purposes, we adopted beta distributed priors with shape parameters 1 and 9, which correspond to a distribution with a mean infection prevalence of 0.10 skewed towards lower values. Values in that range are generally consistent with test positivity for the state of Indiana (41).

### Markov chain Monte Carlo algorithm

To avoid the challenges associated with calculating Pr(***n***) directly, we approximated the posterior distribution of ***θ*** using Markov chain Monte Carlo. Specifically, we used the Metropolis-Hastings algorithm as implemented with default settings in the BayesianTools (42) package in R (43). We ran a total of 100,000 iterations across nine chains, applying a burnin at 10,000 iterations for each and thinning every 100 samples. We assessed convergence through visual inspection of traceplots and calculation of Gelman-Rubin statistics (Fig. S3). We assessed parameter non-identifiability through pairwise correlation plots (Fig. S4).

### Estimation of time-varying prevalence

To obtain an estimate of time-varying prevalence, *Prev*(*t*), among the campus population, we first estimated daily incidence of infection, *I*_*S*_(*t*), from the time series of symptomatic case notifications, as described in (26). To do this, we deconvolved the symptomatic case notifications with the incubation period distribution and the delay from symptom onset to testing. The incubation period was modeled as a log-normal distribution with parameters *μ* = 1. 621 and *σ* = 0.418 (28), and the delay from symptom onset to testing as a Poisson distribution with a mean of two days. We used the backprojNP function in the R surveillance package (version 1.18.0) for the deconvolution (44). We then estimated the total number of infections by date of infection, *I*(*t*) = *I* _*S*_ (*t*) / 0. 57, by assuming that 57% of infections were symptomatic (27) and that all symptomatic infections were ultimately tested under the intense on-campus testing environment. Finally, we estimated *Prev*(*t*) as

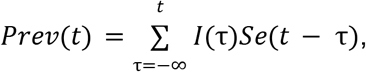

where *Se*(*t*) is an estimate of sensitivity by day of infection by Grassly et al. (11).

### Model validation

Overall, 98% of the 800 simulated parameter values (8 parameters x 100 simulated data sets) fell within their respective 95% credible intervals. For all individual parameters, 95% or more of simulated values fell within their 95% credible intervals. These results suggest that our posterior estimates provide an appropriate description of uncertainty about the true values of the parameters that we sought to estimate. In addition, median values from the posterior distribution were well correlated with simulated values. Pearson correlations ranged from 0.40 for saliva sensitivity to 0.76 for saliva specificity (Fig. S2). Within the range of simulated parameter values we considered, these results suggest that the inference method produces median estimates in the right general direction but that true parameters may lie elsewhere within the credible interval.

## SUPPLEMENTAL TABLES

**Table S1.** Test positivity stratified by test type (columns) and reason participants were tested (rows).

|  | **Commercial** | **Saliva** | **Antigen** |
| --- | --- | --- | --- |
| **Surveillance positivity** | 1.2% (10/802) | 2.5% (20/805) | 0% (0/3) |
| **Non-surveillance positivity** | 6.5% (2/31) | 14.6% (6/41) | 14.7% (5/34) |

**Table S2.**
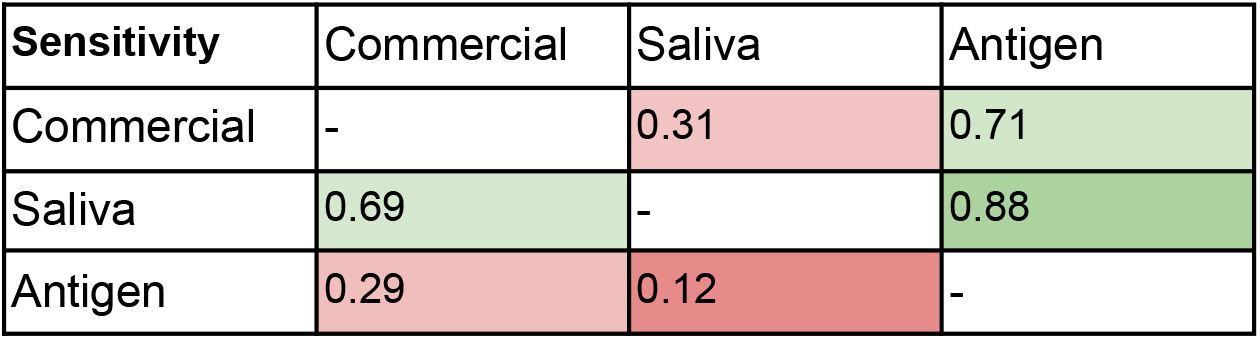
Pairwise probabilities that one type of test (row) is more sensitive than another (column).

**Table S3.** Pairwise probabilities that one type of test (row) is more specific than another (column).

| **Specificity** | Commercial | Saliva | Antigen |
| --- | --- | --- | --- |
| Commercial | - | 0.86 | 0.92 |
| Saliva | 0.14 | - | 0.81 |
| Antigen | 0.08 | 0.19 | - |

## SUPPLEMENTAL FIGURES

**Figure S1.**
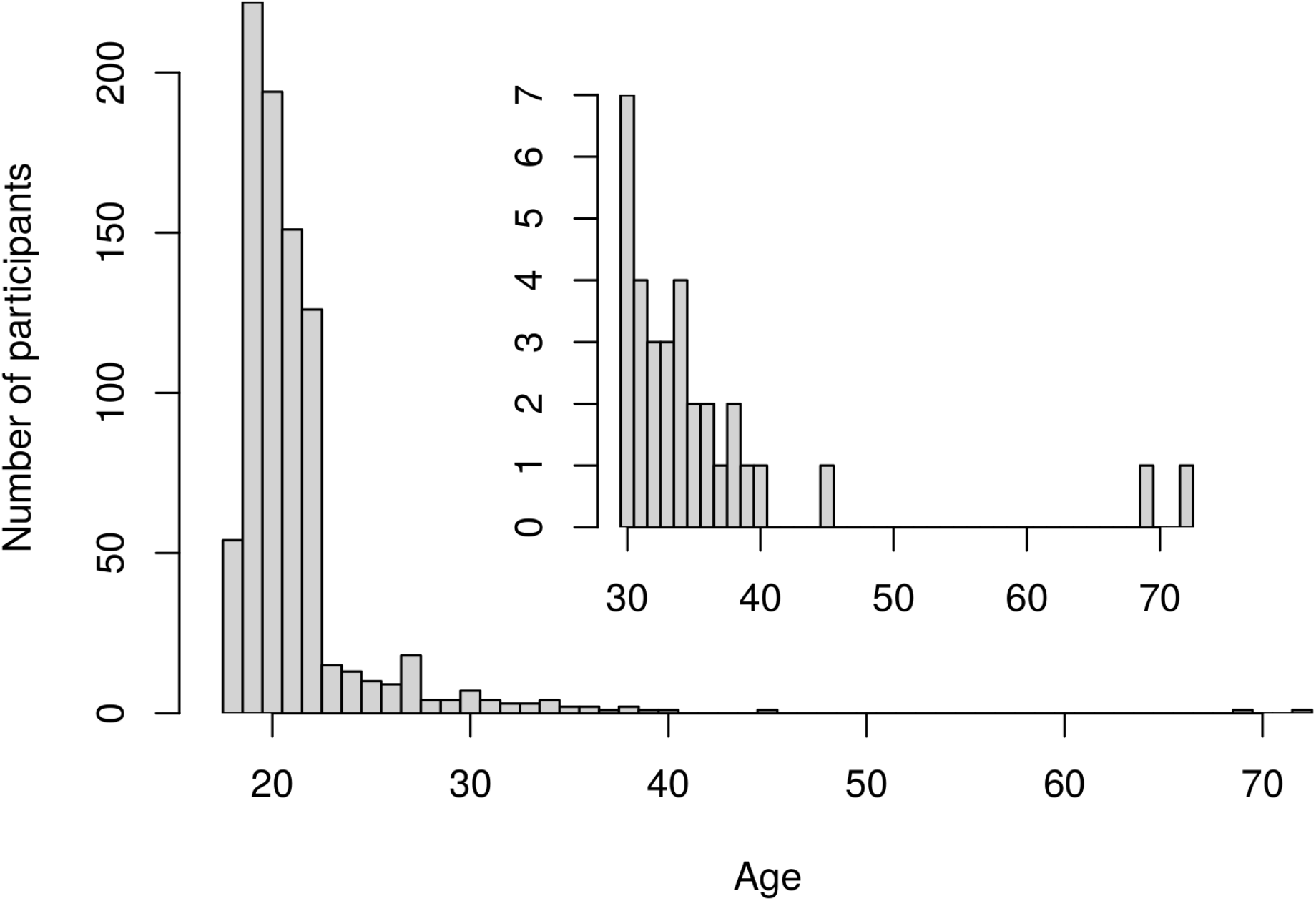
Age distribution of participants. The main panel shows the overall age distribution of study participants, while the inset panel shows the age distribution for individuals 30 and older.

**Figure S2.**
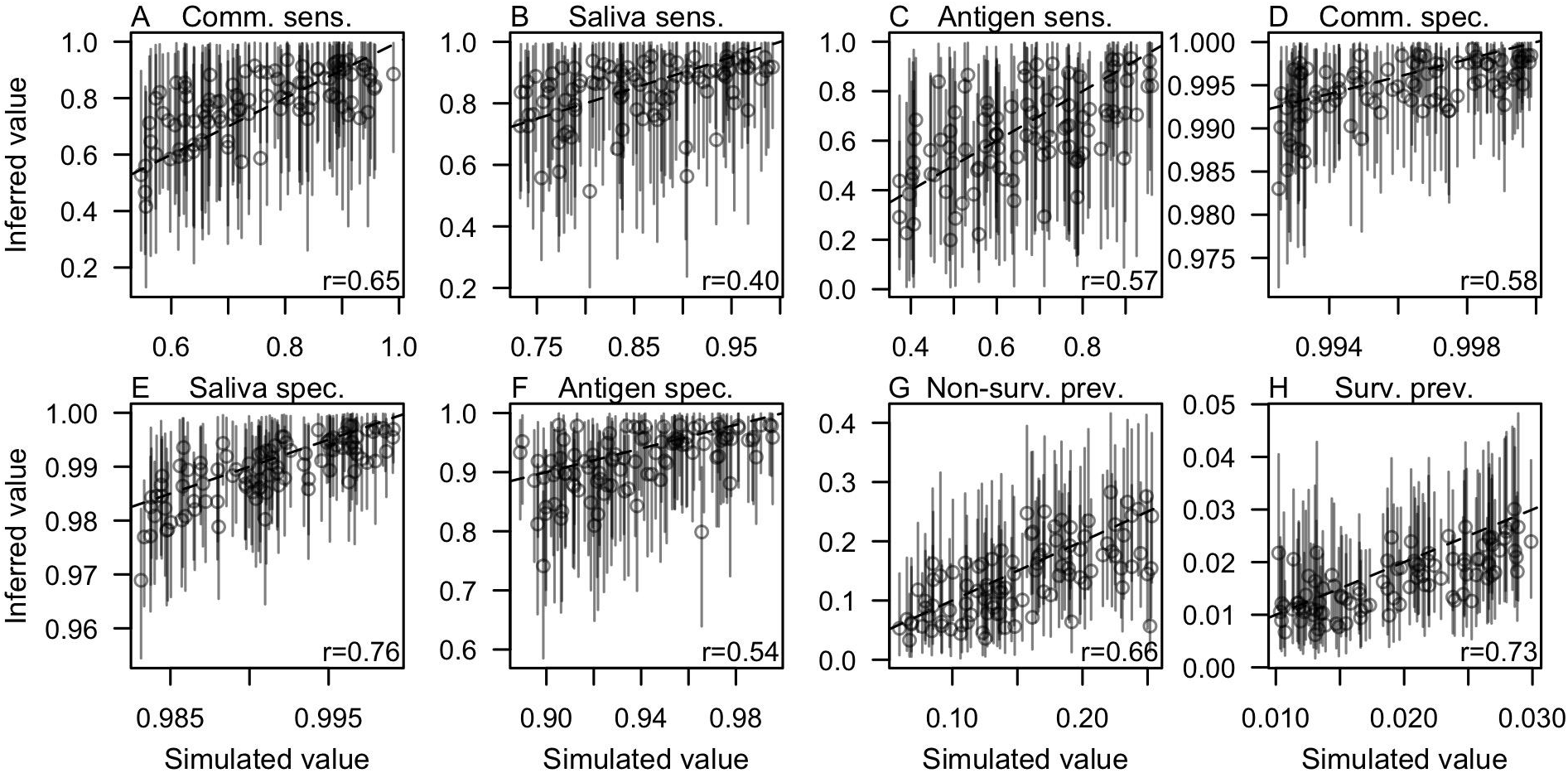
Validation on simulated data. For each of 100 parameter sets drawn uniformly and independently from the 95% credible interval of each parameter (x-axis), we simulated testing outcomes for each individual from each category (i.e., reason for testing, which tests were applied) in our empirical data set. For each of those simulated data sets, we obtained posterior estimates of the eight parameters, the median and 95% credible intervals of which are displayed here (y-axis). The dashed line shows a one-to-one relationship, and the Pearson correlation between simulated and median inferred values for each parameter is displayed in each panel.

**Figure S3.**
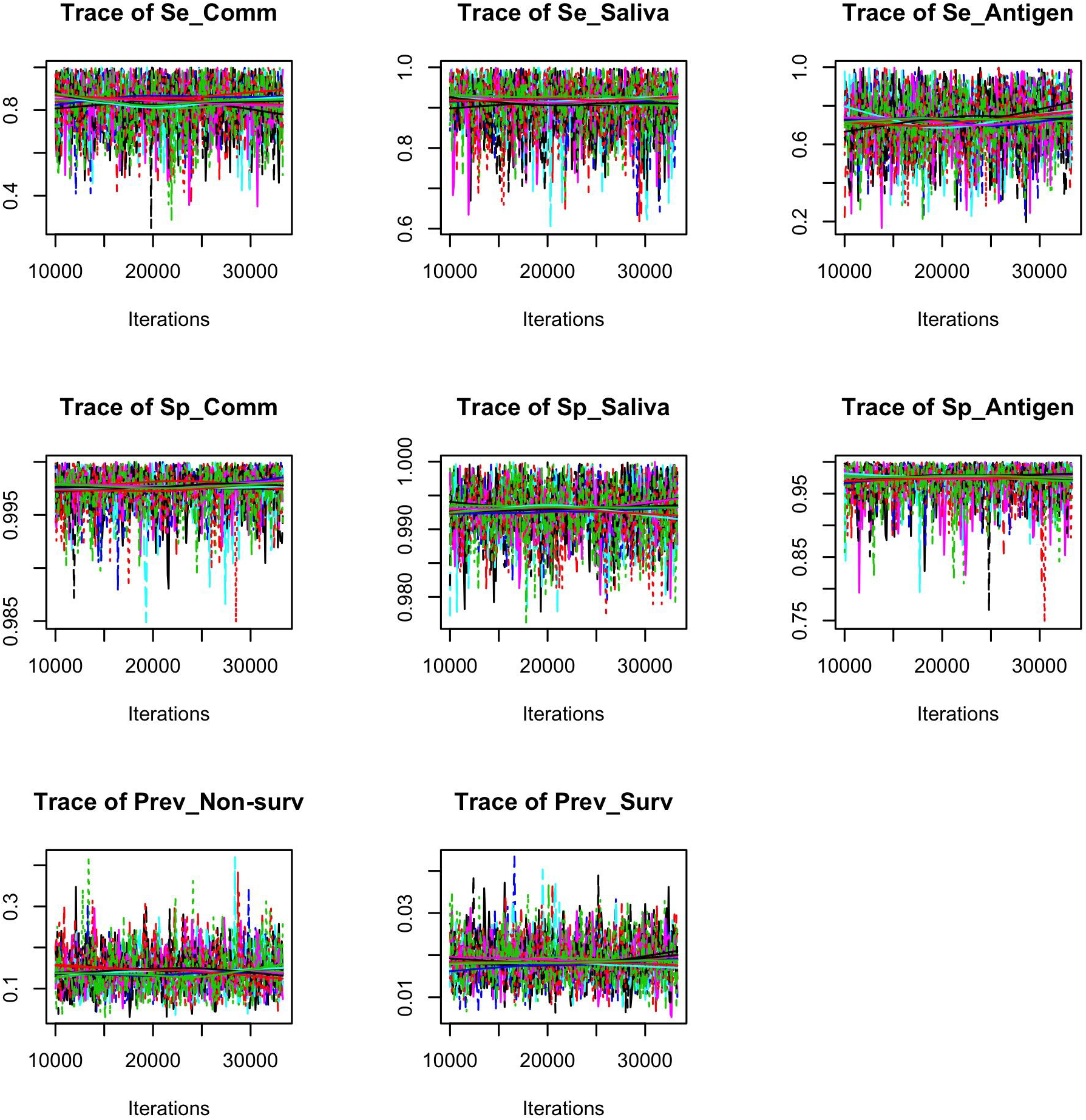
Trace plots of the eight parameters. Traces of the model parameters over the course of the MCMC chains reveal no visual indication of autocorrelation. This interpretation is supported by potential scale reduction factors being at or below 1.01 for all parameters (both for point estimates and upper confidence intervals) and the multivariate potential scale reduction factor being 1.01, as assessed by the gelmanDiagnostics function from the BayesianTools package in R.

**Figure S4.**
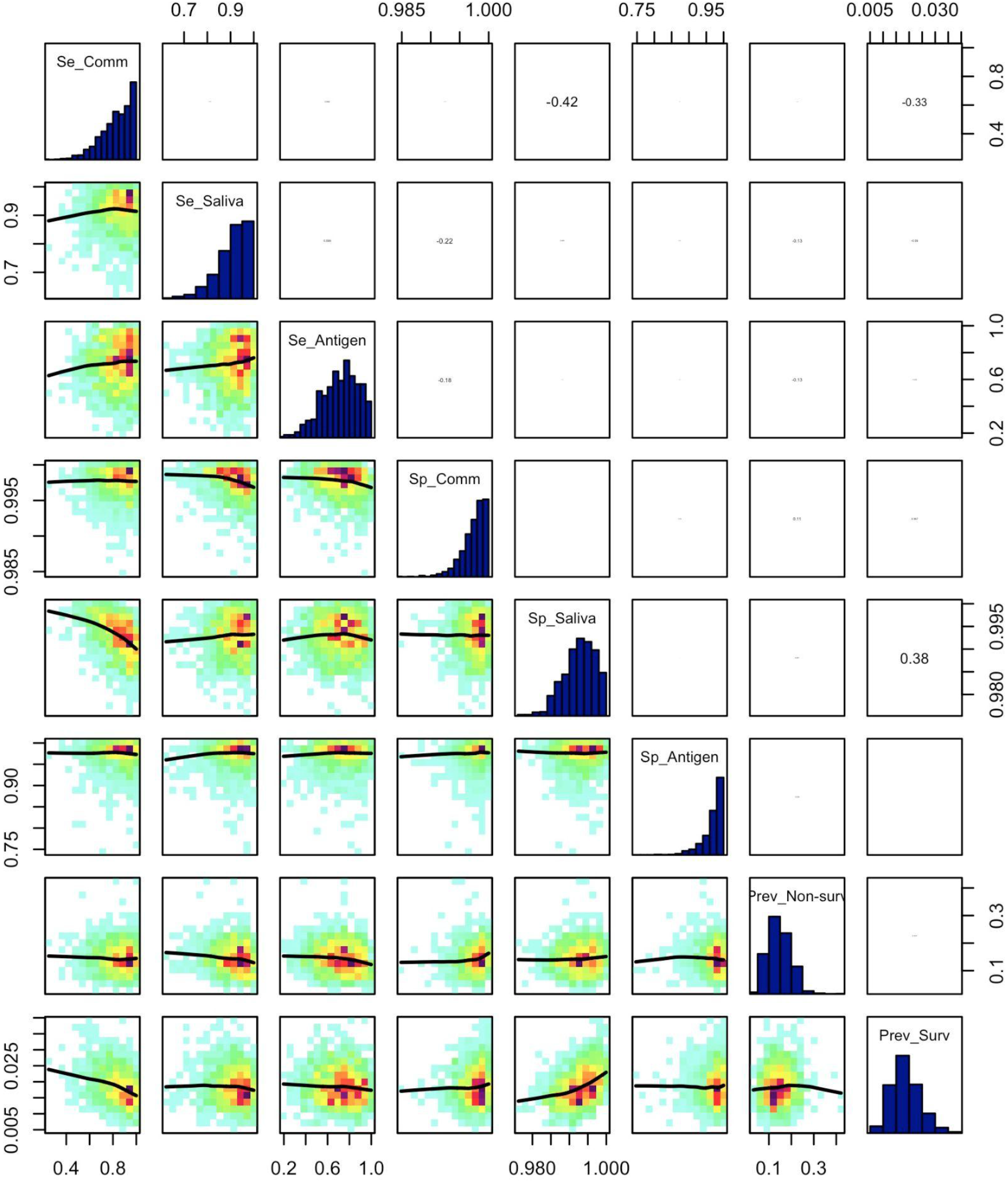
Correlation plot for the eight parameters. The most prominent correlations occurred among *Se*_*Comm*_, *Sp*_*Saliva*_, and *Prev*_*Surv*_. This indicates some discrepancy between commercial and saliva tests about samples collected through surveillance. Specifically, this pattern is consistent with samples with a positive result under the saliva test but a negative result under the commercial test. The existence of these correlations indicates some degree of difficulty in determining the true latent state for samples for which that inconsistency occurs. This figure was made with the correlationPlot function from the BayesianTools library in R.

## REFERENCES

1. Aleta A, Martín-Corral D, Pastore Y Piontti A, Ajelli M, Litvinova M, Chinazzi M, Dean NE, Halloran ME, Longini IM Jr, Merler S, Pentland A, Vespignani A, Moro E, Moreno Y. 2020. Modelling the impact of testing, contact tracing and household quarantine on second waves of COVID-19. Nat Hum Behav 4:964–971.

2. Ng T-C, Cheng H-Y, Chang H-H, Liu C-C, Yang C-C, Jian S-W, Liu D-P, Cohen T, Lin H-H. 2021. Comparison of Estimated Effectiveness of Case-Based and Population-Based Interventions on COVID-19 Containment in Taiwan. JAMA Intern Med https://doi.org/10.1001/jamainternmed.2021.1644.

3. Chin ET, Huynh BQ, Chapman LAC, Murrill M, Basu S, Lo NC. 2020. Frequency of Routine Testing for Coronavirus Disease 2019 (COVID-19) in High-risk Healthcare Environments to Reduce Outbreaks. Clin Infect Dis https://doi.org/10.1093/cid/ciaa1383.

4. Evans S, Agnew E, Vynnycky E, Stimson J, Bhattacharya A, Rooney C, Warne B, Robotham J. 2021. The impact of testing and infection prevention and control strategies on within-hospital transmission dynamics of COVID-19 in English hospitals. Philos Trans R Soc Lond B Biol Sci 376:20200268.

5. VanderWaal K, Black L, Hodge J, Bedada A, Dee S. 2021. Modeling transmission dynamics and effectiveness of worker screening programs for SARS-CoV-2 in pork processing plants. bioRxiv. medRxiv.

6. Wells CR, Townsend JP, Pandey A, Moghadas SM, Krieger G, Singer B, McDonald RH, Fitzpatrick MC, Galvani AP. 2021. Optimal COVID-19 quarantine and testing strategies. Nat Commun 12:356.

7. Panovska-Griffiths J, Kerr CC, Stuart RM, Mistry D, Klein DJ, Viner RM, Bonell C. 2020. Determining the optimal strategy for reopening schools, the impact of test and trace interventions, and the risk of occurrence of a second COVID-19 epidemic wave in the UK: a modelling study. Lancet Child Adolesc Health 4:817–827.

8. Paltiel AD, Zheng A, Walensky RP. 2020. Assessment of SARS-CoV-2 Screening Strategies to Permit the Safe Reopening of College Campuses in the United States. JAMA Netw Open 3:e2016818.

9. Gostic K, Gomez AC, Mummah RO, Kucharski AJ, Lloyd-Smith JO. 2020. Estimated effectiveness of symptom and risk screening to prevent the spread of COVID-19. Elife 9.

10. Kiang MV, Chin ET, Huynh BQ, Chapman LAC, Rodríguez-Barraquer I, Greenhouse B, Rutherford GW, Bibbins-Domingo K, Havlir D, Basu S, Lo NC. 2021. Routine asymptomatic testing strategies for airline travel during the COVID-19 pandemic: a simulation study. Lancet Infect Dis https://doi.org/10.1016/S1473-3099(21)00134-1.

11. Grassly NC, Pons-Salort M, Parker EPK, White PJ, Ferguson NM, Imperial College COVID-19 Response Team. 2020. Comparison of molecular testing strategies for COVID-19 control: a mathematical modelling study. Lancet Infect Dis 20:1381–1389.

12. Larremore DB, Wilder B, Lester E, Shehata S, Burke JM, Hay JA, Tambe M, Mina MJ, Parker R. 2021. Test sensitivity is secondary to frequency and turnaround time for COVID-19 screening. Sci Adv 7.

13. Watson J, Whiting PF, Brush JE. 2020. Interpreting a covid-19 test result. BMJ 369:m1808.

14. Arevalo-Rodriguez I, Buitrago-Garcia D, Simancas-Racines D, Zambrano-Achig P, Del Campo R, Ciapponi A, Sued O, Martinez-García L, Rutjes AW, Low N, Bossuyt PM, Perez-Molina JA, Zamora J. 2020. False-negative results of initial RT-PCR assays for COVID-19: A systematic review. PLoS One 15:e0242958.

15. Wang W, Xu Y, Gao R, Lu R, Han K, Wu G, Tan W. 2020. Detection of SARS-CoV-2 in Different Types of Clinical Specimens. JAMA 323:1843–1844.

16. Wölfel R, Corman VM, Guggemos W, Seilmaier M, Zange S, Müller MA, Niemeyer D, Jones TC, Vollmar P, Rothe C, Hoelscher M, Bleicker T, Brünink S, Schneider J, Ehmann R, Zwirglmaier K, Drosten C, Wendtner C. 2020. Virological assessment of hospitalized patients with COVID-2019. Nature 581:465–469.

17. Kucirka LM, Lauer SA, Laeyendecker O, Boon D, Lessler J. 2020. Variation in False-Negative Rate of Reverse Transcriptase Polymerase Chain Reaction-Based SARS-CoV-2 Tests by Time Since Exposure. Ann Intern Med 173:262–267.

18. Wikramaratna PS, Paton RS, Ghafari M, Lourenço J. 2020. Estimating the false-negative test probability of SARS-CoV-2 by RT-PCR. Euro Surveill 25.

19. Chung E, Chow EJ, Wilcox NC, Burstein R, Brandstetter E, Han PD, Fay K, Pfau B, Adler A, Lacombe K, Lockwood CM, Uyeki TM, Shendure J, Duchin JS, Rieder MJ, Nickerson DA, Boeckh M, Famulare M, Hughes JP, Starita LM, Bedford T, Englund JA, Chu HY. 2021. Comparison of Symptoms and RNA Levels in Children and Adults With SARS-CoV-2 Infection in the Community Setting. JAMA Pediatr https://doi.org/10.1001/jamapediatrics.2021.2025.

20. Dramé M, Tabue Teguo M, Proye E, Hequet F, Hentzien M, Kanagaratnam L, Godaert L. 2020. Should RT-PCR be considered a gold standard in the diagnosis of COVID-19? J Med Virol 92:2312–2313.

21. Rindskopf D, Rindskopf W. 1986. The value of latent class analysis in medical diagnosis. Stat Med 5:21–27.

22. Kostoulas P, Eusebi P, Hartnack S. 2020. Diagnostic accuracy estimates for COVID-19 RT-PCR and Lateral flow immunoassay tests with Bayesian latent class models. Research Square. Research Square.

23. Butler-Laporte G, Lawandi A, Schiller I, Yao M, Dendukuri N, McDonald EG, Lee TC. 2021. Comparison of Saliva and Nasopharyngeal Swab Nucleic Acid Amplification Testing for Detection of SARS-CoV-2: A Systematic Review and Meta-analysis. JAMA Intern Med 181:353–360.

24. Hartnack S, Eusebi P, Kostoulas P. 2021. Bayesian latent class models to estimate diagnostic test accuracies of COVID-19 tests. J Med Virol.

25. Ranoa DRE, Holland RL, Alnaji FG, Green KJ, Wang L, Brooke CB, Burke MD, Fan TM, Hergenrother PJ. 2020. Saliva-Based Molecular Testing for SARS-CoV-2 that Bypasses RNA Extraction. bioRxiv.

26. Cavany S, Bivins A, Wu Z, North D, Bibby K, Perkins TA. 2021. Inferring SARS-CoV-2 RNA shedding into wastewater relative to time of infection. medRxiv.

27. Kasper MR, Geibe JR, Sears CL, Riegodedios AJ, Luse T, Von Thun AM, McGinnis MB, Olson N, Houskamp D, Fenequito R, Burgess TH, Armstrong AW, DeLong G, Hawkins RJ, Gillingham BL. 2020. An Outbreak of Covid-19 on an Aircraft Carrier. N Engl J Med 383:2417–2426.

28. Lauer SA, Grantz KH, Bi Q, Jones FK, Zheng Q, Meredith HR, Azman AS, Reich NG, Lessler J. 2020. The Incubation Period of Coronavirus Disease 2019 (COVID-19) From Publicly Reported Confirmed Cases: Estimation and Application. Ann Intern Med 172:577–582.

29. Wyllie AL, Fournier J, Casanovas-Massana A, Campbell M, Tokuyama M, Vijayakumar P, Warren JL, Geng B, Muenker MC, Moore AJ, Vogels CBF, Petrone ME, Ott IM, Lu P, Venkataraman A, Lu-Culligan A, Klein J, Earnest R, Simonov M, Datta R, Handoko R, Naushad N, Sewanan LR, Valdez J, White EB, Lapidus S, Kalinich CC, Jiang X, Kim DJ, Kudo E, Linehan M, Mao T, Moriyama M, Oh JE, Park A, Silva J, Song E, Takahashi T, Taura M, Weizman O-E, Wong P, Yang Y, Bermejo S, Odio CD, Omer SB, Dela Cruz CS, Farhadian S, Martinello RA, Iwasaki A, Grubaugh ND, Ko AI. 2020. Saliva or Nasopharyngeal Swab Specimens for Detection of SARS-CoV-2. N Engl J Med 383:1283–1286.

30. Hanson KE, Barker AP, Hillyard DR, Gilmore N, Barrett JW, Orlandi RR, Shakir SM. 2020. Self-Collected Anterior Nasal and Saliva Specimens versus Health Care Worker-Collected Nasopharyngeal Swabs for the Molecular Detection of SARS-CoV-2. J Clin Microbiol 58.

31. Teo AKJ, Choudhury Y, Tan IB, Cher CY, Chew SH, Wan ZY, Cheng LTE, Oon LLE, Tan MH, Chan KS, Hsu LY. 2021. Saliva is more sensitive than nasopharyngeal or nasal swabs for diagnosis of asymptomatic and mild COVID-19 infection. Sci Rep 11:3134.

32. Lee RA, Herigon JC, Benedetti A, Pollock NR, Denkinger CM. 2021. Performance of Saliva, Oropharyngeal Swabs, and Nasal Swabs for SARS-CoV-2 Molecular Detection: a Systematic Review and Meta-analysis. J Clin Microbiol 59.

33. Pray IW. 2021. Performance of an antigen-based test for asymptomatic and symptomatic SARS-CoV-2 testing at two university campuses—Wisconsin, September--October 2020. MMWR Morb Mortal Wkly Rep 69.

34. Corman VM, Haage VC, Bleicker T, Schmidt ML, Mühlemann B, Zuchowski M, Jo WK, Tscheak P, Möncke-Buchner E, Müller MA, Krumbholz A, Drexler JF, Drosten C. 2021. Comparison of seven commercial SARS-CoV-2 rapid point-of-care antigen tests: a single-centre laboratory evaluation study. Lancet Microbe 2:e311–e319.

35. Mina MJ, Parker R, Larremore DB. 2020. Rethinking Covid-19 Test Sensitivity - A Strategy for Containment. N Engl J Med 383:e120.

36. He X, Lau EHY, Wu P, Deng X, Wang J, Hao X, Lau YC, Wong JY, Guan Y, Tan X, Mo X, Chen Y, Liao B, Chen W, Hu F, Zhang Q, Zhong M, Wu Y, Zhao L, Zhang F, Cowling BJ, Li F, Leung GM. 2020. Temporal dynamics in viral shedding and transmissibility of COVID-19. Nat Med 26:672–675.

37. Christensen H, Turner K, Trickey A, Booton RD, Hemani G, Nixon E, Relton C, Danon L, Hickman M, Brooks-Pollock E, Part of the University of Bristol UNCOVER group. 2020. COVID-19 transmission in a university setting: a rapid review of modelling studies. bioRxiv. medRxiv.

38. Hamer DH, White LF, Jenkins HE, Gill CJ, Landsberg HE, Klapperich C, Bulekova K, Platt J, Decarie L, Gilmore W, Pilkington M, MacDowell TL, Faria MA, Densmore D, Landaverde L, Li W, Rose T, Burgay SP, Miller C, Doucette-Stamm L, Lockard K, Elmore K, Schroeder T, Zaia AM, Kolaczyk ED, Waters G, Brown RA. 2021. Assessment of a COVID-19 Control Plan on an Urban University Campus During a Second Wave of the Pandemic. JAMA Netw Open 4:e2116425.

39. Motta FC, McGoff KA, Deckard A, Wolfe CR, Moody MA, Cavanaugh K, Denny TN, Harer J, Haase SB. 2021. Benefits of surveillance testing and quarantine in a SARS-CoV-2 vaccinated population of students on a university campus. bioRxiv. medRxiv.

40. Pei S, Yamana TK, Kandula S, Galanti M, Shaman J. 2021. Overall burden and characteristics of COVID-19 in the United States during 2020. bioRxiv. medRxiv.

41. 2021. ISDH - Novel Coronavirus: Indiana COVID-19 Dashboard and Map.

42. Hartig F, Minunno F, Paul S. 2018. BayesianTools: General-Purpose MCMC and SMC Samplers and Tools for Bayesian Statistics, R package version 0.1. 3.

43. R Core Team. 2018. R: A Language and Environment for Statistical Computing. R Foundation for Statistical Computing, Vienna, Austria.

44. Meyer S, Held L, Höhle M. 2017. Spatio-Temporal Analysis of Epidemic Phenomena Using the R Package surveillance. Journal of Statistical Software.

